# Application of Social Vulnerability Index to Identify High- risk Population of Contracting COVID-19 Infection: a state-level study

**DOI:** 10.1101/2020.08.03.20166983

**Authors:** Odalys Estefania Lara-Garcia, Violeta Alvarez Retamales, Oswaldo Madrid Suarez, Priyanka Parajuli, Susan Hingle, Robert Robinson

## Abstract

Social factors that determine a population’s health are known as the social determinants of health. During the past few weeks, as COVID-19 cases grew exponentially, the discrepancy among the number of cases distribution was evident.By applying the social vulnerability index and analyzing data from a total of 102 counties across the state of Illinois, we investigated which factors enhanced the risk of contracting the infection and which were related to a lower risk of infection. Our results showed that social factors such as belonging to a minority group, speaking English less than well, living in a multi-unit structure, and households with individuals of age group of 17 or younger were associated with a higher risk of infection. On the other hand, we found that factors such as living in a mobile home, individuals of age group 65 or older, low income per capita and, older than age 5 with disability were protective. We propose that communities with disproportionate health burdens can be identified by the application of these factors. Future efforts need to focus on decreasing the gap of to decrease the gap of disparity by modifying these social factors.

## Introduction

Social determinants of health (SDOH) are everyday life factors able to impact a population’s health, by defining the conditions in which a person lives, works, and grows (1). These factors include socioeconomic status, housing type, access to healthcare, employment, disabilities, and minority status. Populations that belong to lower socioeconomic status are inherently linked to worse health outcomes (1). A tool to measure the applicability of these factors has been developed by the CDC to aid public health officials in identifying communities with the greatest need for support. Social Vulnerability Index(SVI) takes into consideration SDOH factors and correlates them with the endurance of communities when faced with a disease outbreak or other threats to human health.

In the past few weeks, as population data has been gathered in the midst of the COVID-19 pandemic, it has become evident that there are some disparities in groups within the same population. More specifically, reported cases of COVID-19 appeared to differ depending on the zip code. An evidence of this has been the distribution of cases among Illinois counties. However, limited information has been available to evaluate the social characteristics that correlate with a higher rate of infection in these communities. In this study, we aim to describe the strength of correlation of social vulnerability index factors and how these play a role in putting the Illinois population at higher risk of becoming infected with COVID-19.

## Methods

Population data for each county in Illinois were obtained from the 2018 American Community Survey from the US Census Bureau. (U.S. Census Bureau; American Community Survey, 2018). The rate of COVID-19 cases per 100,000 people was calculated for each county using data from the Illinois Department of Public Health website on April 27, 2020 (https://www.dph.illinois.gov/covid19/covid19-statistics).

SVI data was obtained from the CDC for each county in Illinois. Data for all SVI themes and theme components were downloaded and used for analysis. The themes and components of the SVI are outlined in Table 1. The percentile rank (EPL) for each component was used for correlation analysis. Higher percentile ranks indicate a greater burden of social vulnerability in a county.

**Table 1.** CDC SVI Themes and components

| Theme | Component |
| --- | --- |
| **Socioeconomic status** | Below poverty level<br>Unemployed<br>Per capita income<br>No high school diploma |
| **Household composition and disability** | Age 65 or older<br>Age 17 or younger<br>Older than age 5 with a disability<br>Single parent households |
| **Minority status and language** | Minority<br>Speaks English “less than well” |
| **Housing type and transportation** | Multi-unit structures<br>Mobile homes<br>Crowding<br>No vehicle<br>Group quarters |

Spearman correlations between the number of COVID-19 cases per 100,000 people and the components of the SVI were determined.

Statistical analyses were performed using SPSS version 25 (SPSS Inc., Chicago, IL, USA). A p-value of 0.05 was chosen for statistical significance.

Maps were prepared using Tableau Desktop Professional Edition, version 2020.1.3 (Tableau Software, LLC).

Centers for Disease Control and Prevention/ Agency for Toxic Substances and Disease Registry/ Geospatial Research, Analysis, and Services Program. Social Vulnerability Index 2018 Database: Illinois. data-and-tools-download.html. Accessed on May 1, 2020.

## Results

All 102 counties in Illinois were included in our analysis. At the time of our evaluation, the counties with the greatest number of cases per capita included Cook, Lake, DuPage, Kankakee, Will, Schuyler,Jasper, Rock Island, Warren, Stark,Tazewell, Saline,Pulaski and, Randolph (FIGURE 1.) Factors that increase the risk of infection that had a statistically significant correlation coefficient included belonging to a minority group(r= 0.4,p <0.001), speaking English less than well(r=0.3, p<0.002), multi unit structure (r=0.3,p=0.001), age group of 17 or younger(r=0.18,p=0.05). Other factors with a negative correlation that were found to be protective were mobile homes (r=-0.34,p<0.001), age group 65 or older(r=-0.3, p=0.002), low income per capita (r=-0.21, p=0.029) and, older than age 5 with disability (r=-0.27, p=0.005).(Table 2)

**Table 2.** SVI component with correlation coefficients

| SVI Theme | SVI Component | Correlation Coefficient | p-Value |
| --- | --- | --- | --- |
| **Socioeconomic Status** | Below Poverty Level | -0.041 | 0.680 |
|  | Unemployed | 0.001 | 0.993 |
|  | Low Per Capita Income | -0.216 | 0.029 |
|  | No High School Diploma | 0.054 | 0.592 |
| **Household Composition and Disability** | Age 65 or Older | -0.300 | 0.002 |
|  | Age 17 or Younger | 0.188 | 0.058 |
|  | Older than Age 5 with Disability | -0.277 | 0.005 |
|  | Single Parent Household | 0.113 | 0.258 |
| **Minority Status And Language** | Minority | 0.410 | <0.001 |
|  | Speaks English "Less than Well" | 0.353 | <0.002 |
| **Housing Type and Transportation** | Multi-Unit Structures | 0.312 | 0.001 |
|  | Mobile Homes | -0.341 | <0.001 |
|  | Crowding | 0.108 | 0.278 |
|  | No Vehicle | -0.066 | 0.510 |
|  | Group Quarters | -0.018 | 0.857 |

## Discussion

Of the nearly 120,000 cases of COVID-19 in Illinois, there were clear variations in the rate of infection among counties based on social factors. Minority group status, English proficiency, multi-unit structures increased the risk of contracting the infection, while owning a mobile home, low income per capita,households of individuals age 65 or older and those with a disability above the age of 5 were protective factors.

Racial and ethnic minority groups have been disproportionately impacted during this pandemic (2). We found that belonging to a minority had a moderate correlation with an increased risk for contracting COVID-19. Wadhera et al, in a recent analysis of the city of New York by borough, found that these factors correlated with the disparities among the distribution of COVID-19 cases and related deaths. Boroughs that had the highest representation of minorities also had a lower socioeconomic status compared to those in which minorities were underrepresented and had a higher rate of cases and associated deaths(2)..A few hypotheses which may be able to explain this phenomenon are based on the social factors that define the conditions in which minorities live in “pre-pandemic” states. Routine access to health care, health literacy, socioeconomic status, and neighborhood environment all play a role in determining an individual’s health outcome. Minorities are more likely to belong to a low socioeconomic group, have lower health literacy, and less access to health care. Minorities are more likely to be “essential workers”, such as bus drivers, grocery clerks; who cannot afford to work from home and are routinely in contact with the public (3). These groups, in addition to having a high risk for COVID-19 infection, also carry a greater risk of having a severe course of the disease. The highest burden of chronic diseases including hypertension and diabetes are found among minority groups such as African Americans and Latinos(4,5). The case and death rate disparity by zip code can be attributed to the population represented. In areas in which Black Americans and Hispanics are overrepresented, there are striking differences in cases and associated deaths. For example, on the South Side of Chicago, where most of the Illinois COVID-19 deaths are concentrated, the population is predominantly African American and Hispanic. (6)

We also found lack of English proficiency was related to a higher risk of infection.We theorize that individuals who belong to groups that “speak English less than well” are likely to have less access to culturally sensitive and high-quality public health information. Evidence of the disproportionate impact on the Hispanic community can be found when analyzing the demographic distribution of cases in counties that comprise the city of Chicago. Latinos by far had a greater rate of COVID-19 cases compared to all other ethnic groups (7).

Household composition also contributes to the risk of infection. Residents of multi-unit structures physically share common areas with others and may have a higher difficulty practicing social distance. The opposite is true for mobile home residents, who can afford a smoother transition to social distancing compared to individuals living in multi-unit structures, as at baseline they do not share common ground with others. Households with individuals older than age 5 with disabilities had a lower risk of infection. This might be explained by the fact that these individuals are less likely to be employed according to a report by the US Bureau of Labor Statistics and as a consequence have a bigger opportunity of practicing social distancing. (8). A similar protective effect was found for households with individuals age 65 or older, and may be assisted by younger family members in doing their groceries and shopping for them. Given the age group we hypothesise that these individuals are more likely to be retired thus increasing the ease of staying at home. An unforeseen result we obtained was that low income per capita was associated with lower risk. We propose that such results may be driven by the fact that individuals from low socioeconomic status are less likely to be tested therefore deceivingly appearing to have less cases associated with this factor.

## Conclusion

Social determinants of health are part of a complex equation that despite tremendous heterogeneity within the same population, is able to predict the health outcome of individuals. Communities with disproportionate health burdens can be identified by the application of these factors. The purpose of this manuscript is to share understanding of the social factors that determine vulnerability and susceptibility to disease in a community, with the ultimate intention that future efforts to decrease the gap of disparity are targeted to the modification of these factors.

## Data Availability

Population data has been obtained from the U.S Census Bureau

## Annex

**Table 3.**
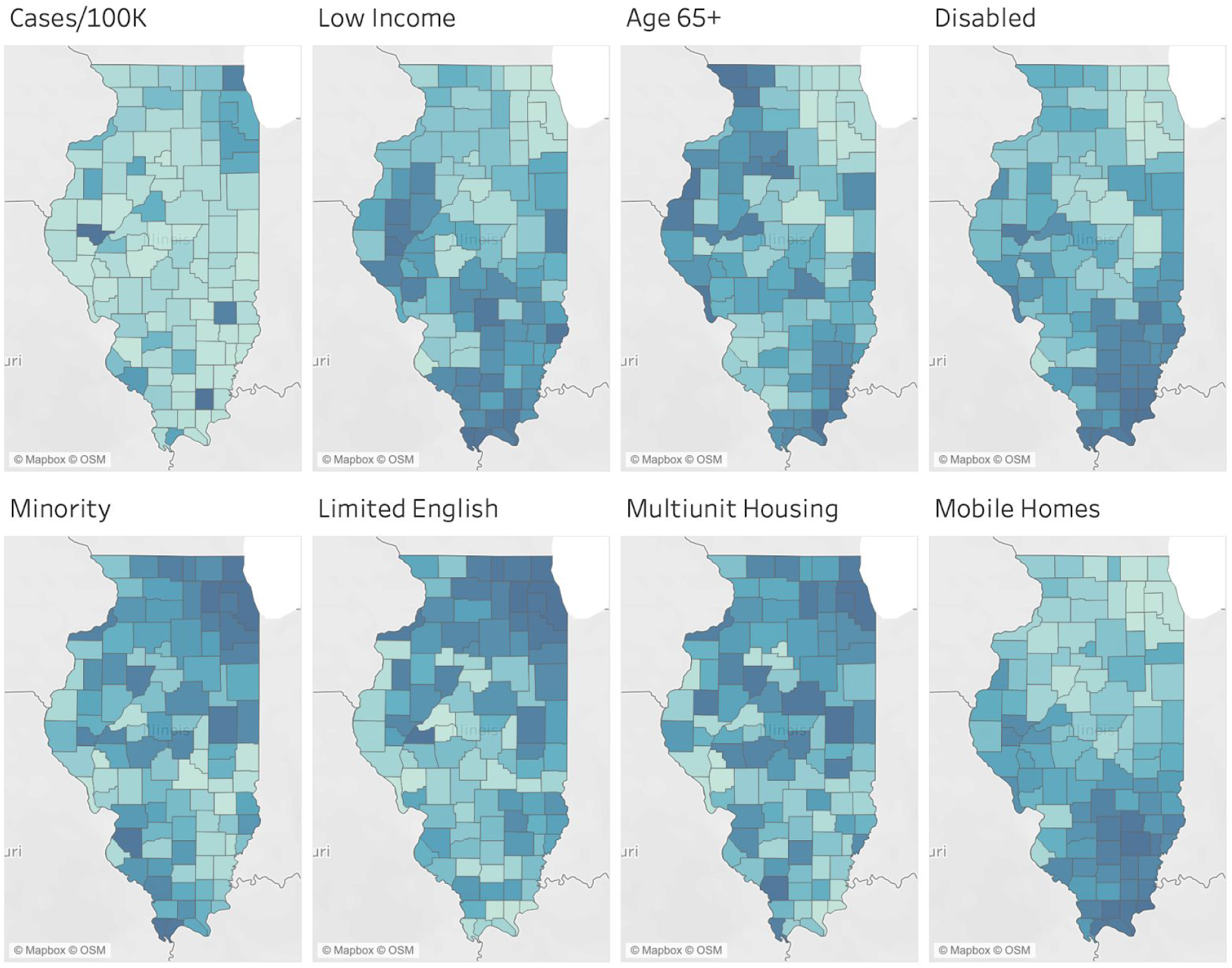
Graphical COVID-19 cases and SVI component representation by Illinois counties.

**Table 4.**
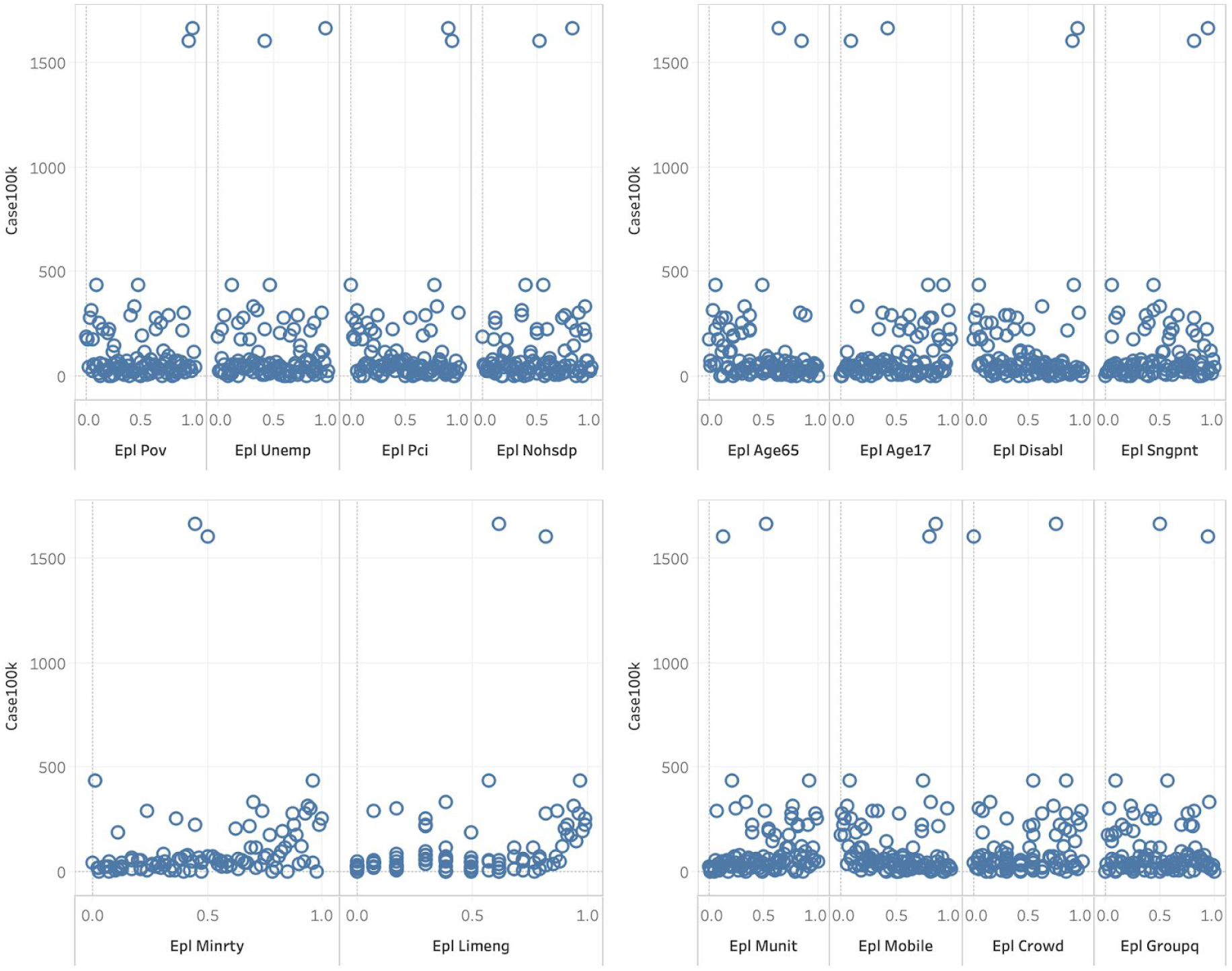
Correlation coefficient graph

